# Dynamic changes in plasma sphingosine-1-phosphate concentrations and mortality in sepsis: a prospective cohort study in the emergency department

**DOI:** 10.64898/2026.11.03.26367220

**Authors:** Binh Van Tran, Ching-Lu Chen, Thang Ngoc Nguyen, You-Cheng Lin, Kai-Chun Hu, Chin-Wang Hsu, Shih-Chang Hsu

## Abstract

**Background:** Sepsis is a dynamic process with evolving host responses, biomarker changes, and organ dysfunction. Sphingosine-1-phosphate (S1P) is a bioactive lipid involved in endothelial and immune regulation. However, the clinical relevance of dynamic S1P changes in sepsis remains unclear. This study aimed to assess their association with mortality.

**Methods:** We conducted a prospective cohort study of adults recruited from the emergency department of Wan Fang Hospital between April 2022 and October 2024. Plasma S1P was measured by ELISA on days 1, 3, and before discharge or death. The primary outcome was 60-day mortality. S1P trajectories were defined by baseline concentrations and longitudinal slopes estimated using linear mixed-effects models. Low-decreasing, low-stable, and high-decreasing trajectories were grouped as descending; the others as ascending. Cox proportional hazards models assessed mortality associations with baseline S1P, trajectory groups, and time-varying S1P.

**Results:** Among 278 participants, 156 had sepsis. Baseline S1P concentrations were lower in patients with sepsis (82.2 [IQR, 47.8–141.1] ng/mL) than in those with non-septic infection (129.2 [IQR, 68.1–306.9] ng/mL) and non-infected controls (132.8 [IQR, 74.1–300.2] ng/mL). In the sepsis cohort, 101 patients were classified into the descending S1P trajectory group and 55 into the ascending group. The descending group had a higher hazard of 60-day mortality (adjusted hazard ratio [aHR] 3.43; 95% confidence interval [CI], 1.11–10.58; p = 0.032). Baseline S1P was not significantly associated with 60-day mortality (aHR 1.004 per 10 ng/mL decrease; 95% CI, 0.977–1.032; p = 0.803). Each 10 ng/mL decrease in time-varying S1P was associated with a 3.5% higher mortality hazard (aHR 1.035; 95% CI, 1.002–1.076; p = 0.041).

**Conclusions:** Declining plasma S1P over time was associated with higher mortality in sepsis. Dynamic changes in S1P may be clinically relevant beyond baseline levels.

## Background

Sepsis remains a major global health burden, with an estimated 21.4 million deaths (31.5% of all deaths worldwide) in 2021 [1]. Sepsis is a dynamic process involving interactions between pathogens and the host immune system, with evolving pro- and anti-inflammatory responses [2]. Septic shock, a severe form of sepsis, is characterized by acute circulatory failure and profound cellular and metabolic abnormalities resulting from a dysregulated host response to infection [3]. Endothelial dysfunction contributes to impaired tissue perfusion, inadequate oxygen delivery, and subsequent multiple organ dysfunction [3]. Therefore, reliable indicators are essential for early identification, prognostic assessment, and management of sepsis [4, 5].

The Sequential Organ Failure Assessment (SOFA) score is widely used to assess organ dysfunction and evaluate prognosis in sepsis [6]. However, its calculation requires multiple clinical and laboratory parameters, potentially limiting its utility for real-time monitoring. Several circulating biomarkers, including C-reactive protein (CRP), procalcitonin (PCT), lactate, and interleukin-6 (IL-6), have been extensively investigated for their diagnostic and prognostic value [7]. Nevertheless, given the complex and dynamic nature of sepsis, no single biomarker can fully capture its pathophysiological processes. Consequently, the discovery of novel biomarkers remains a research priority in sepsis.

Sphingosine-1-phosphate (S1P) is a bioactive sphingolipid mediator constitutively produced and released by endothelial cells and erythrocytes [8–10]. S1P is involved in various biological processes, including vascular homeostasis, endothelial barrier integrity, immune cell trafficking, and inflammatory responses [11]. In circulation, approximately 60% of S1P is carried by apolipoprotein M (ApoM)-containing high-density lipoprotein (HDL) and 30% by albumin, its two major plasma carriers [12, 13]. Extracellular S1P mediates its biological effects through five G protein-coupled receptors (S1PR1–5), with S1PR1 highly expressed on endothelial cells [14]. Given its role in sepsis pathophysiology, S1P may serve as a promising prognostic biomarker.

Despite consistent evidence that circulating S1P concentrations are reduced in patients with sepsis, its prognostic value for mortality remains uncertain [15–17]. In addition, most studies have focused on baseline S1P, which may not capture the evolving pathophysiology of sepsis [16, 18, 19]. Furthermore, changes in S1P sources and carrier proteins during sepsis may affect its circulating concentrations [17, 20, 21]. These fluctuations may reflect changes in sepsis severity and progression. We therefore hypothesized that longitudinal S1P measurements may provide additional prognostic information beyond baseline levels. Accordingly, this study aimed to investigate the association between changes in plasma S1P concentrations over time and 60-day mortality in patients with sepsis.

## Methods

### Participants

This observational, single-center, prospective cohort study was conducted at Taipei Municipal Wanfang Hospital (Taipei, Taiwan) between April 2022 and October 2024. Adults (≥ 18 years) presenting to the emergency department (ED) with suspected sepsis (qSOFA ≥ 2) and non-infected controls were consecutively enrolled. Patients with infection were classified as sepsis or non-septic infection according to the sepsis-3 criteria [6]. Exclusion criteria included severe immunosuppression, active malignancy, pregnancy, and breastfeeding. These conditions were excluded because they may independently influence S1P metabolism and signaling, immune responses, and clinical outcomes [22, 23]. The clinical study was approved by the Taipei Medical University Institutional Review Board (TMU-JIRB No. N202202022). Written informed consent was obtained from all participants or from their legally authorized representatives (Fig. 1).

**Fig. 1.**
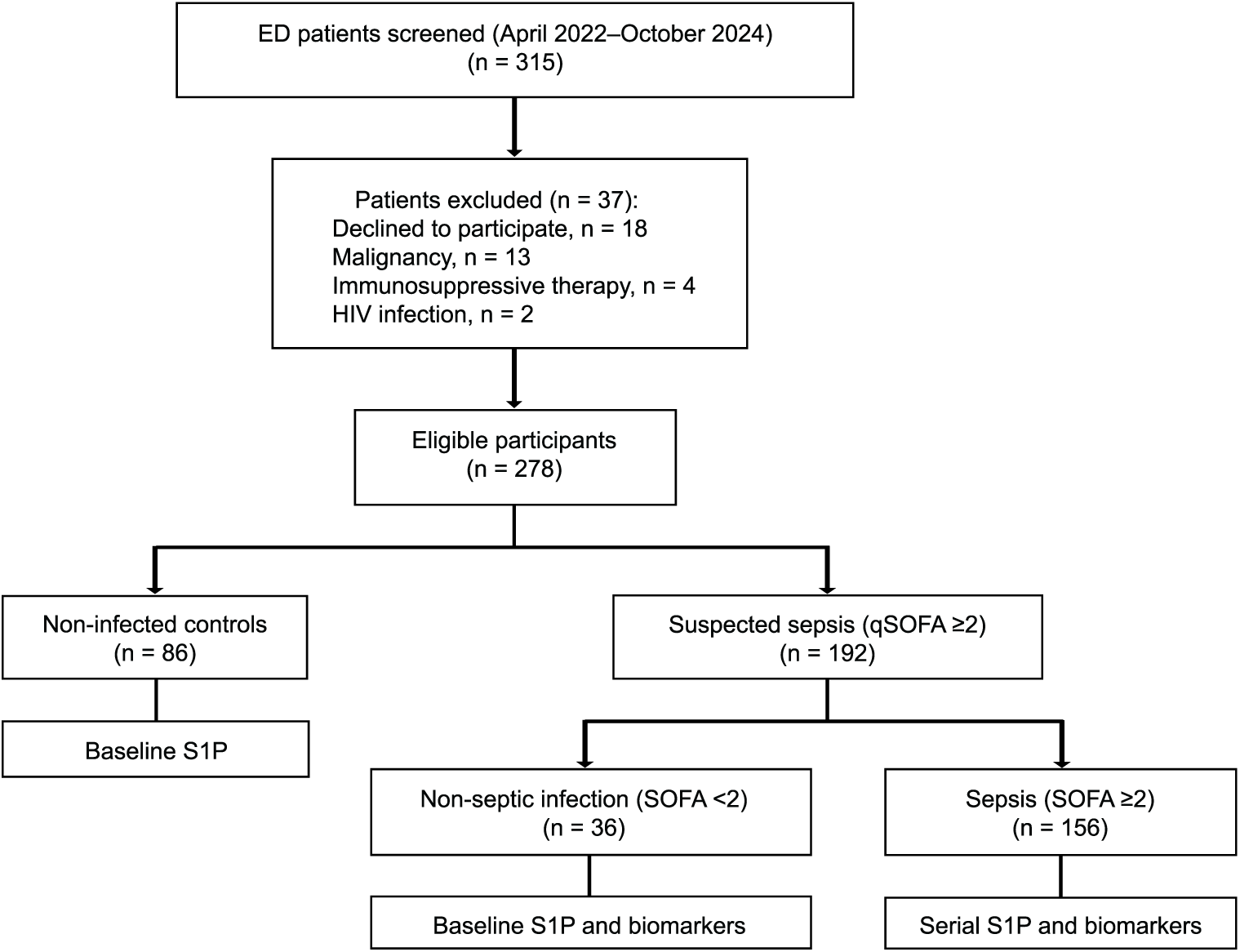
STROBE flow diagram of participant enrollment and study procedures. Abbreviations: ED, emergency department; HIV, human immunodeficiency virus; S1P, sphingosine-1-phosphate; qSOFA, quick Sequential Organ Failure Assessment; SOFA, Sequential Organ Failure Assessment.

### S1P measurement

Trained research nurses collected peripheral venous blood samples (10 mL) at predefined time points. Baseline samples (T0) were collected from all participants upon ED presentation. In patients with sepsis, additional samples were obtained on day 3 of hospitalization (T1) and one day before hospital discharge or death (T2). The collected blood samples were stored in EDTA-containing tubes and immediately centrifuged at 2,500 × g for 10 minutes. The resulting plasma was stored at −80°C until analysis. Plasma S1P and IL-6 concentrations were measured using commercially available enzyme-linked immunosorbent assay (ELISA) kits from MyBioSource and BioLegend, respectively.

### Data collection

Demographic data including age, sex, body weight, body mass index (BMI), and comorbidities were collected for all participants. Detailed clinical and laboratory data were obtained only in patients with suspected infection (non-septic infection and sepsis groups). Clinical parameters recorded at ED presentation included Glasgow Coma Scale (GCS) score, heart rate, mean arterial pressure (MAP), respiratory rate, peripheral oxygen saturation (SpO_2_), body temperature, and urine output. The primary infection source was categorized as respiratory tract, intra-abdominal, urinary tract, soft tissue, or other. Laboratory data included hematological parameters (red blood cell count, hemoglobin, white blood cell count, neutrophil count, and platelet count), biochemical parameters (creatinine, lactate, bilirubin, glucose, albumin, and HDL), inflammatory markers (CRP and PCT), and blood culture results. Patients with suspected infection were managed according to the Surviving Sepsis Campaign (SSC) 2021 guidelines [24].

### Outcome assessment

The primary outcome was 60-day all-cause mortality, defined as death from any cause within 60 days of ED admission. Time-to-event was defined as the interval from ED admission to death or censoring. Patients alive at 60 days were administratively censored at 60 days, whereas those lost to follow-up were censored at their last known alive date. Secondary outcomes included intensive care unit (ICU) admission, and hospital length of stay.

### Statistical analysis

Before formal analysis, normality of continuous variables was assessed using the Shapiro–Wilk test. Normally distributed variables were presented as mean with standard deviation (SD) and compared using the independent-samples t-test or one-way ANOVA, as appropriate. Otherwise, variables were presented as medians with interquartile ranges (IQRs) and compared using the Mann–Whitney U test or the Kruskal–Wallis test, as appropriate. Categorical variables were expressed as counts with percentages and compared using the Chi-square or Fisher’s exact test.

Patients with sepsis were stratified into high- and low-baseline S1P groups using the median S1P concentration of non-infected controls as the cutoff. A linear mixed-effects model with random intercepts and slopes was used to model the longitudinal change in S1P and estimate individual slopes. Patients were then classified into decreasing, stable, or increasing groups according to the 25th and 75th percentiles of the estimated slope distribution. Six S1P trajectory categories were defined: high-decreasing, high-stable, high-increasing, low-decreasing, low-stable, and low-increasing. Based on a prespecified hypothesis that low baseline S1P and decreasing trajectories would be associated with higher mortality risk, low-decreasing, low-stable, and high-decreasing categories were grouped as descending. In contrast, the remaining three categories were grouped as ascending.

Kaplan–Meier survival curves were constructed to illustrate survival probabilities in the ascending and descending S1P trajectory groups, with comparisons performed using the log-rank test. Univariable Cox regression analyses were used to estimate hazard ratios (HRs) and 95% confidence intervals (CIs) for variables associated with mortality in previous studies. Covariates with missing data less than 20% and p values < 0.20 in univariable Cox regression were included in the multivariable Cox regression model. To minimize baseline imbalance between the ascending and descending S1P trajectory groups, propensity score matching was performed using 1:1 nearest-neighbor matching. Covariate balance was assessed using standardized mean differences (SMDs), with an SMD < 0.1 indicating adequate balance. Multivariable Cox regression was subsequently performed in the matched cohort. A time-varying Cox proportional hazards model was additionally performed to account for temporal changes in S1P exposure. To ensure model validity, the proportional hazards assumption, autocorrelation, and homoscedasticity were assessed, with the linearity of quantitative variables previously verified.

Statistical analyses were conducted using R statistical software (version 4.4.2, https://www.r-project.org). Linear mixed-effects models, survival analyses, and propensity score matching were performed using the “lme4”, “survival”, and “MatchIt” packages, respectively. A two-sided p-value < 0.05 was considered statistically significant.

## Results

### Baseline characteristics of the study population

The study included 278 participants: 86 non-infected controls, 36 patients with non-septic infections, and 156 patients with sepsis. Non-infected controls were younger (48.2 ± 20.5 years) and had a lower proportion of males (36.0%) than the other groups (both p < 0.05). Age, sex, and BMI were comparable between the non-septic infection and sepsis groups (all p > 0.05). However, the Charlson Comorbidity Index (CCI) was significantly higher in patients with sepsis than in those with non-septic infections (2.6 ± 1.8 versus 1.8 ± 1.6; p < 0.05) (Table 1). Compared with patients with non-septic infections, those with sepsis had significantly higher CRP levels (11.4 [IQR, 5.0–18.3] versus 2.5 [IQR, 0.5–7.2] mg/L; p < 0.001) and SOFA scores (3.40 ± 2.03 versus 1.27 ± 1.55; p < 0.001) (Supplementary Table S1).

**Table 1.**
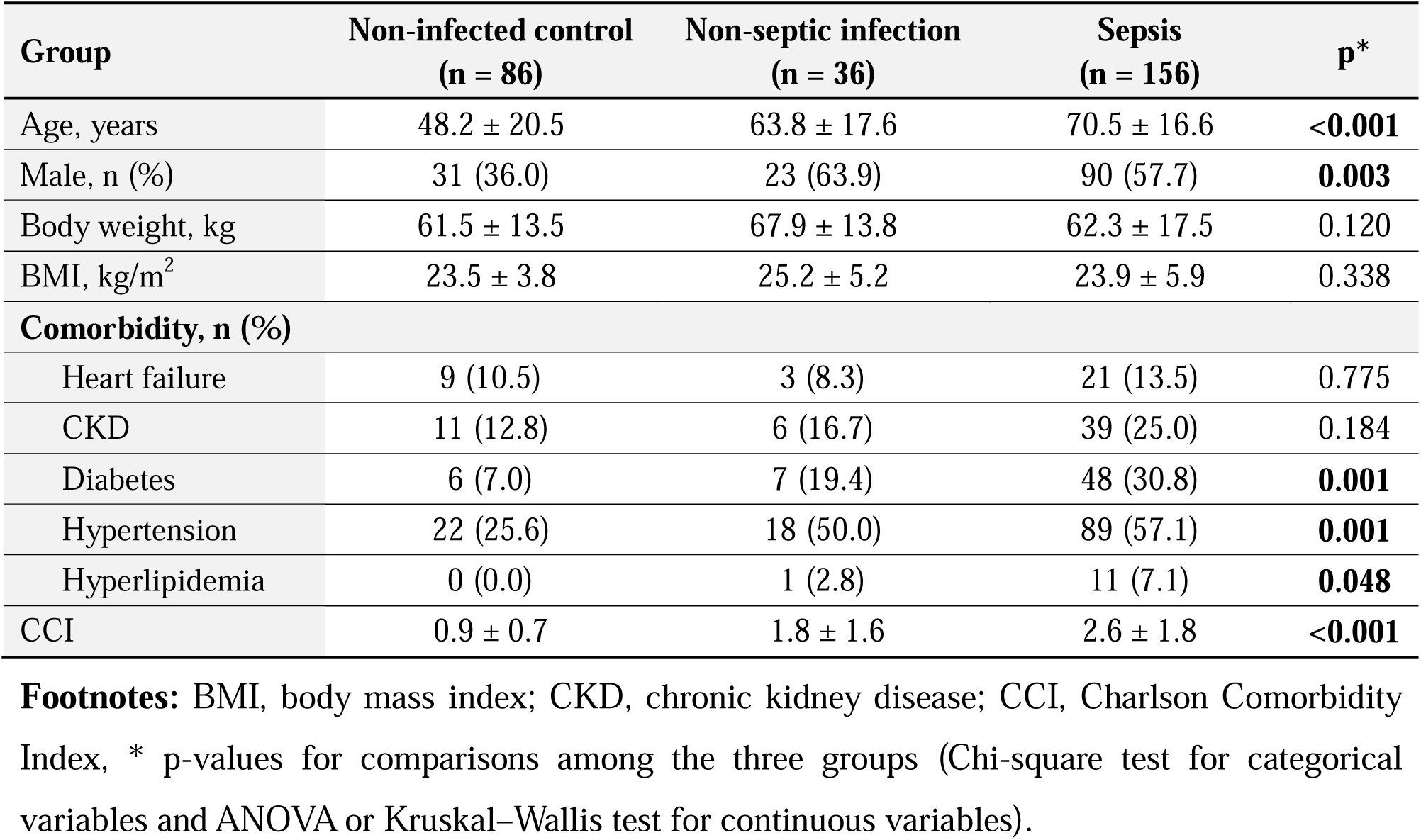
Baseline characteristics of study groups.

Baseline plasma S1P concentrations were significantly lower in patients with sepsis (82.2 [IQR, 47.8–141.1] ng/mL) than in non-infected controls (132.8 [IQR, 74.1–300.2] ng/mL; p < 0.001) and patients with non-septic infection (129.2 [IQR, 68.1–306.9] ng/mL; p < 0.05) (Fig. 2). Within the sepsis cohort, those with septic shock had significantly higher SOFA scores (5.46 ± 2.77 versus 2.99 ± 1.57; p < 0.001) and baseline IL-6 concentrations (237.0 [IQR, 126.5–653.9] versus 83.6 [IQR, 21.1–261.3] pg/mL; p < 0.001) than those without shock. However, baseline S1P concentrations did not differ significantly between the two groups (79.0 [IQR, 38.4–128.4] versus 82.5 [IQR, 51.8–144.5] ng/mL; p > 0.05) (Supplementary Table S2). Correlation analysis showed a weak positive association between S1P and IL-6 (r = 0.250; p < 0.01) and a weak negative association with SOFA scores (r = −0.166; p < 0.05) (Supplementary Table S3).

**Fig. 2.**
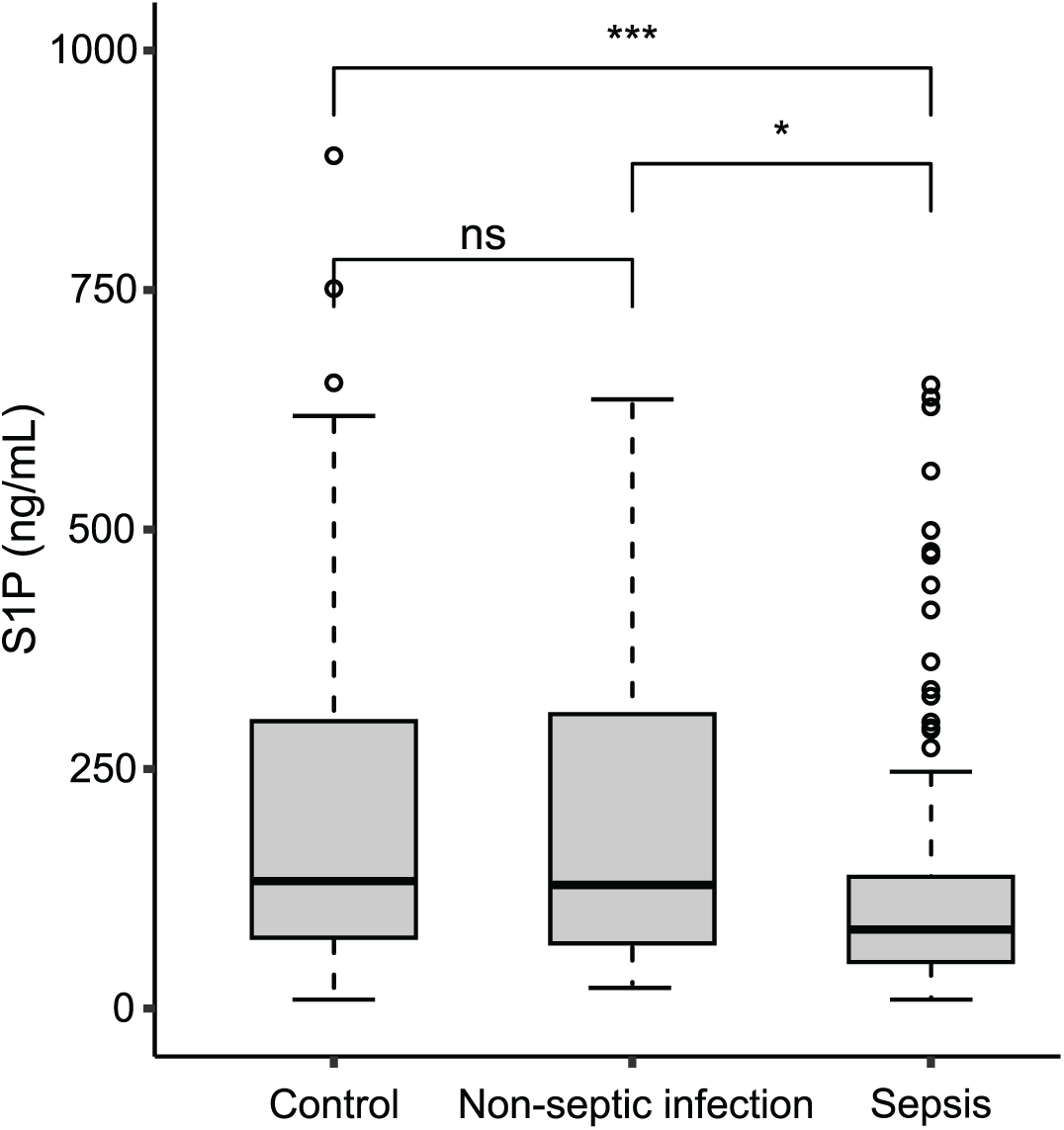
Distribution of baseline plasma S1P concentrations in the non-infected control, non-septic infection, and sepsis groups. Abbreviations: S1P, sphingosine-1-phosphate; ns, non-significant; * p < 0.05; *** p < 0.001.

### S1P trajectory classification

Six S1P trajectory categories were identified based on the baseline cutoff of 132.8 ng/mL derived from non-infected controls and individual S1P slopes. The low-stable category was the most common (65 patients; 41.7%), whereas the high-decreasing category was the least frequent (6 patients; 3.8%). Compared with the high-increasing category, the low-decreasing, low-stable, and high-decreasing categories had adjusted HRs greater than 1, with the highest estimate observed in the high-decreasing (adjusted HR 10.48; 95% CI, 1.90–57.89) (Fig. 3). For further analysis, the six categories were divided into two predefined trajectory groups: descending (n = 101) and ascending (n = 55).

**Fig. 3.**
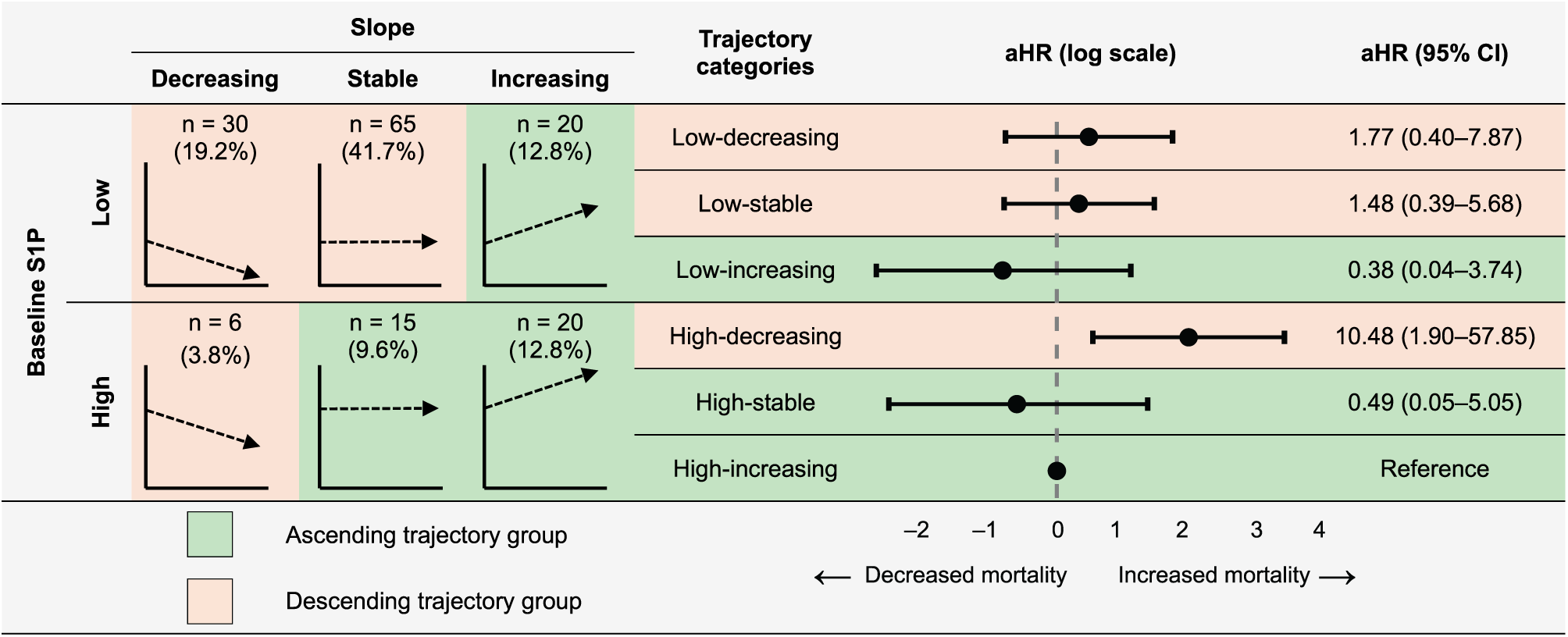
Distribution of S1P trajectory categories and their associations with 60-day mortality. Footnotes: Baseline S1P concentrations were stratified as high or low using the median value of the non-infected control group (132.8 ng/mL). S1P trajectories were characterized using slopes derived from longitudinal S1P measurements. Based on the slope distribution, patients were categorized into decreasing (<25th percentile), stable (25th–75th percentile), and increasing (>75th percentile) trajectories.

### Baseline characteristics by S1P trajectory groups

The ascending and descending groups showed no significant differences in age, sex, BMI, comorbidities, CCI, other clinical and laboratory parameters, or SOFA scores (all p > 0.05). Baseline S1P concentrations were 161.0 [IQR, 71.0–295.7] ng/mL in the ascending group and 70.5 [IQR, 44.2–97.0] ng/mL in the descending group (p < 0.05). ICU admission rates (9.1% versus 12.9%) and hospital length of stay (11.0 [IQR, 8.0–17.0] versus 10.0 [IQR, 8.0–13.0] days) did not differ significantly between groups (both p > 0.05). The 60-day mortality rate was 9.1% (5/55) in the ascending group versus 16.8% (17/101) in the descending group (p > 0.05) (Table 2).

**Table 2.** Baseline characteristics stratified by S1P trajectory groups.

|  | **Overall<br>(n = 156)** | **Ascending<br>(n = 55)** | **Descending<br>(n = 101)** | **p** |
|---|---|---|---|---|
| Age, years | 70.5 ± 16.6 | 73.4 ± 16.8 | 68.9 ± 16.3 | 0.117 |
| Male, n (%) | 90 (57.7) | 32 (58.2) | 58 (57.4) | 1.000 |
| Body weight, kg | 62.3 ± 17.5 | 61.4 ± 16.6 | 62.8 ± 18.01 | 0.653 |
| **Comorbidity, n (%)** |  |  |  |  |
| Heart failure | 21 (13.5) | 10 (18.2) | 11 (10.9) | 0.303 |
| CKD | 39 (25.0) | 13 (23.6) | 26 (25.7) | 0.923 |
| Diabetes | 48 (30.8) | 19 (34.5) | 29 (28.7) | 0.567 |
| Hypertension | 88 (56.4) | 32 (58.2) | 56 (55.4) | 0.873 |
| Hyperlipidemia | 11 (7.1) | 3 (5.5) | 8 (7.9) | 0.804 |
| CCI, points | 2.6 ± 1.8 | 2.5 ± 1.5 | 2.7 ± 2.0 | 0.439 |
| **Source of infection, n (%)** |  |  |  |  |
| Respiratory tract | 69 (44.2) | 22 (40.0) | 47 (46.5) | 0.501 |
| Intra-abdominal | 20 (12.8) | 8 (14.5) | 12 (11.9) | 0.625 |
| Urinary tract | 36 (23.1) | 16 (29.1) | 20 (19.8) | 0.233 |
| Soft tissue | 24 (15.4) | 9 (16.4) | 15 (14.9) | 0.819 |
| Others/Unknown | 7 (4.5) | 0 (0.0) | 7 (6.9) | 0.052 |
| **Clinical signs** |  |  |  |  |
| Heart rate, bpm | 98.9 ± 24.9 | 99.4 ± 22.7 | 98.7 ± 26.1 | 0.858 |
| MAP, mmHg | 95.2 ± 20.8 | 93.8 ± 20.3 | 95.9 ± 21.1 | 0.535 |
| Respiratory rate, bpm | 21.1 ± 5.5 | 20.5 ± 4.6 | 21.4 ± 5.9 | 0.318 |
| Temperature, ºC | 37.5 ± 1.2 | 37.5 ± 1.1 | 37.4 ± 1.2 | 0.595 |
| SpO<sub>2</sub>, % | 94.4 ± 6.4 | 95.1 ± 4.5 | 94.0 ± 7.3 | 0.302 |
| Urine output, mL/kg/min | 0.54 ± 0.39 | 0.54 ± 0.37 | 0.55 ± 0.39 | 0.861 |
| SOFA score | 3.4 ± 2.0 | 3.2 ± 1.8 | 3.5 ± 2.1 | 0.357 |
| Septic shock, n (%) | 26 (16.7) | 6 (10.9) | 20 (19.8) | 0.230 |
| **Baseline laboratory data** |  |  |  |  |
| Creatinine, mg/dL | 1.1 (0.9–1.6) | 1.1 (0.9–1.6) | 1.1 (0.8–1.8) | 0.664 |
| CRP, mg/L | 11.4 (5.0–18.3) | 10.8 (3.5–17.7) | 11.5 (5.6–18.5) | 0.322 |
| Lactate, mmol/L | 2.0 (1.4–3.1) | 2.1 (1.3–3.1) | 1.9 (1.4–3.1) | 0.785 |
| Albumin, g/L | 3.1 ± 0.5 | 3.0 ± 0.5 | 3.1 ± 0.5 | 0.529 |
| IL-6, pg/mL | 101.5 (29.8–351.6) | 103.0 (38.1–498.9) | 95.7 (27.9–237.9) | 0.253 |
| S1P, ng/mL | 82.0 (48.0–141.0) | 161.0 (71.0–295.7) | 70.5 (44.2–97.0) | **<0.01** |
| Positive blood culture, n (%) | 52 (33.3) | 18 (32.7) | 34 (33.7) | 1.000 |
| **Outcomes** |  |  |  |  |
| ICU admission rate, n (%) | 18 (11.5) | 5 (9.1) | 13 (12.9) | 0.657 |
| Hospital length of stay, days | 10.0 (8.0–14.3) | 11.0 (8.0–17.0) | 10.0 (8.0–13.0) | 0.149 |
| 60-day mortality, n (%) | 22 (14.1) | 5 (9.1) | 17 (16.8) | 0.277 |

**Footnotes:** CKD, chronic kidney disease; CCI, Charlson Comorbidity Index; MAP, mean arterial pressure; SpO<sub>2</sub>, peripheral oxygen saturation; SOFA, Sequential Organ Failure Assessment; CRP, C-reactive protein; IL-6, interleukin-6; S1P, sphingosine-1-phosphate; ICU, intensive care unit.

### S1P trajectory groups and mortality

In univariable Cox regression, the descending group had an HR of 1.94 (95% CI, 0.72–5.26) compared with the ascending group. Kaplan–Meier analysis showed no significant difference in survival between groups (log-rank p > 0.05). After adjustment for age, sex, CCI, SOFA score, and baseline IL-6, the descending group had an adjusted HR of 3.43 (95% CI, 1.11–10.58; p = 0.032). Following propensity score matching, baseline covariates were generally balanced between groups (Supplementary Table S4), and the adjusted HR was 3.61 (95% CI, 1.16–11.25; p = 0.027) (Fig. 4).

**Fig. 4.**
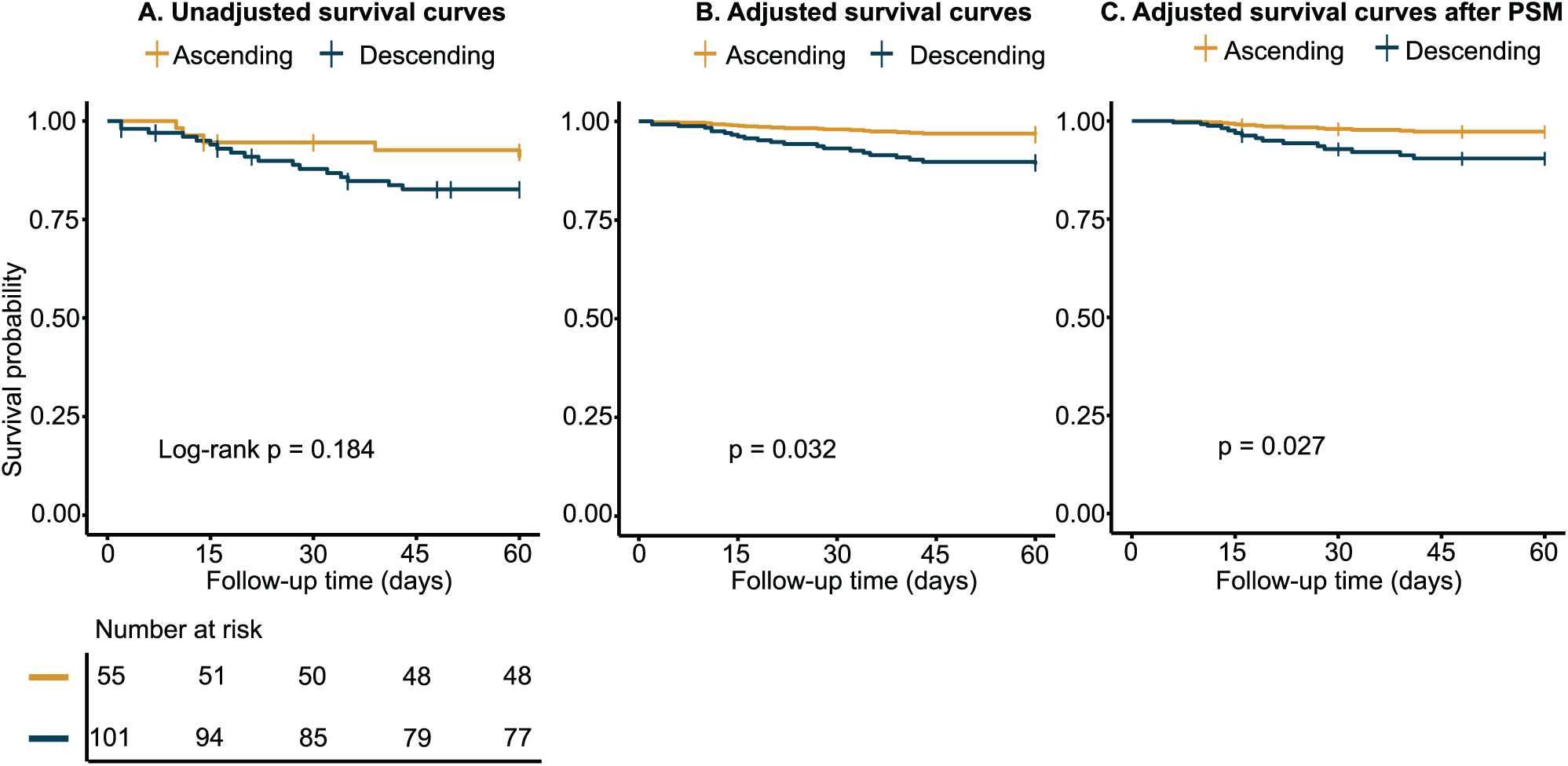
Kaplan–Meier survival curves by S1P trajectory group. (A) Unadjusted Kaplan–Meier survival curves for 60-day mortality. (B) Adjusted survival curves derived from a Cox proportional hazards model adjusted for age, sex, CCI, baseline SOFA score, and baseline IL-6 concentration. (C) Adjusted survival curves after propensity score matching (PSM), with adjustment for age, sex, CCI, baseline SOFA score, and baseline IL-6 concentration. Abbreviations: S1P, sphingosine-1-phosphate; CCI, Charlson Comorbidity Index; SOFA, Sequential Organ Failure Assessment; IL-6, interleukin-6; PSM, propensity score matching.

### Time-varying S1P concentrations and mortality

Baseline S1P was not independently associated with 60-day mortality (adjusted HR 1.004 per 10 ng/mL decrease; 95% CI, 0.977–1.032; p > 0.05). Similarly, univariable time-varying Cox regression showed no significant association (HR 1.010 per 10 ng/mL decrease; 95% CI, 0.980–1.030; p > 0.05). However, after adjustment for age, sex, CCI, SOFA score, and time-varying IL-6, each 10 ng/mL decrease in S1P was associated with a 3.5% higher hazard of 60-day mortality (adjusted HR 1.035; 95% CI, 1.002–1.076; p = 0.041). Older age, higher SOFA scores, and higher IL-6 concentrations were also associated with increased mortality hazards in both baseline and time-varying Cox models (Table 3).

**Table 3.** Comparison of baseline and time-varying Cox regression analyses for factors associated with 60-day mortality.

| **Variable** | **Univariate Cox regression** |  | **Baseline Cox regression<sup>a</sup>** |  | **Time-varying Cox regression<sup>b</sup>** |  |
|---|---|---|---|---|---|---|
|  | **HR (95% CI)** | **p** | **aHR (95% CI)** | **p** | **aHR (95% CI)** | **p** |
| Age, years | 1.053 (1.021–1.086) | **0.001** | 1.045 (1.006–1.085) | **0.022** | 1.057 (1.017–1.099) | **0.005** |
| Male sex | 1.171 (0.506–2.710) | 0.188 | 1.266 (0.489–3.275) | 0.627 | 1.401 (0.544–3.603) | 0.485 |
| Weight, kg | 0.995 (0.968–1.022) | 0.697 | – | – | – | – |
| CCI, points | 1.234 (1.008–1.511) | **0.042** | 1.032 (0.794–1.341) | 0.816 | 0.995 (0.768–1.289) | 0.968 |
| SOFA score, points | 1.386 (1.197–1.607) | **<0.001** | 1.351 (1.138–1.604) | **<0.001** | 1.384 (1.166–1.643) | **<0.001** |
| CRP, mg/L | 0.994 (0.948–1.042) | 0.790 | – | – | – | – |
| Baseline IL-6, per 10 pg/mL increase | 1.006 (1.002–1.010) | **0.001** | 1.006 (1.002–1.011) | **0.010** | – | – |
| Baseline S1P, per 10 ng/mL decrease | 0.985 (0.959–1.012) | 0.277 | 1.004 (0.977–1.032) | 0.803 | – | – |
| Time-varying IL-6, per 10 pg/mL increase | 1.013 (1.008–1.018) | **<0.001** | – | – | 1.020 (1.010–1.030) | **<0.001** |
| Time-varying S1P, per 10 ng/mL decrease | 1.010 (0.980–1.030) | 0.663 | – | – | 1.035 (1.002–1.076) | **0.041** |

**Footnotes:** S1P was analyzed as a continuous variable, with hazard ratios expressed per 10 ng/mL increase in all models. <sup>a</sup>Baseline multivariate Cox regression model including variables with p < 0.20 in the univariate analysis. <sup>b</sup>Time-varying Cox regression model adjusted for age, sex, CCI, SOFA score, and time-varying IL-6. HR, hazard ratio; aHR, adjusted hazard ratio; CI, confident interval; CCI, Charlson Comorbidity Index; SOFA, Sequential Organ Failure Assessment; CRP, C-reactive protein; IL-6, interleukin-6; S1P, sphingosine-1-phosphate.

## Discussion

This single-center prospective cohort study investigated dynamic changes in plasma S1P concentrations in patients with sepsis presenting to the ED. Baseline S1P concentrations were lower in sepsis and weakly inversely correlated with organ dysfunction severity. However, baseline levels alone were not independently associated with mortality. In contrast, longitudinal analyses suggested a relationship between S1P changes over time and mortality. Specifically, the descending trajectory group and lower time-varying S1P concentrations were associated with an increased hazard of 60-day mortality.

Our study found significantly reduced baseline plasma S1P concentrations in patients with sepsis, consistent with previous findings [16, 18, 25]. Notably, S1P levels did not differ significantly between patients with non-septic infection and non-infected controls. Furthermore, baseline S1P concentrations were inversely correlated with sepsis severity, as assessed by SOFA scores [16]. A previous study reported relatively preserved S1P concentrations at lower SOFA scores (≤ 3), with marked reductions at scores ≥ 8 [17]. These findings suggest that S1P concentrations may be maintained in infections not meeting the sepsis-3 criteria [6]. Although age and sex distributions differed significantly between groups, neither significantly influenced S1P levels [16, 26, 27]. Similarly, S1P levels showed no significant correlation with CCI in our cohort. Available evidence suggests that reduced S1P levels may not be fully attributable to underlying comorbidities [28].

Baseline S1P levels were not associated with mortality in our cohort. This finding was supported by both univariable and multivariable Cox regression analyses. Consistent with our results, a substudy of the ALBIOS trial reported no significant differences in 90-day mortality across S1P tertiles [15]. Similarly, another clinical study found no significant difference in S1P levels between 28-day survivors and non-survivors [29]. Growing evidence nevertheless highlights the clinical relevance of S1P in sepsis-related complications. S1P has been identified as a strong indicator of septic shock [16]. In addition, baseline S1P has been shown to independently predict coagulation failure [15]. Furthermore, sphingolipid metabolism has been implicated in endothelial injury and sepsis-induced ARDS [19, 25]. Altered S1P-HDL interactions have also been linked to the need for mechanical ventilation [30]. In contrast, dynamic changes in S1P may provide prognostic insights into mortality in patients with sepsis. In particular, declining S1P levels over time were associated with higher mortality risk. Trajectory and time-varying analyses incorporate updated levels, potentially capturing the relationship between S1P and mortality more effectively [31]. This approach may be more appropriate given the dynamic nature of sepsis, characterized by continuous interactions among multiple mediators, including S1P [2, 32].

Several biological mechanisms may explain the association between declining S1P levels and mortality in our study. S1P has been shown to preserve the endothelial glycocalyx by reducing syndecan-1 shedding [33, 34]. S1P also regulates immune cell trafficking and inflammatory responses, contributing to host defense and disease tolerance during infection [32]. In sepsis, loss of S1P sources and decreased levels of its carrier molecules may contribute to lower circulating concentrations [10, 17]. Excessive fluid resuscitation may further reduce S1P levels through hemodilution [35]. Declining S1P concentrations may contribute to vascular leakage, impaired tissue perfusion, and multiple organ dysfunction [34, 36]. In addition, the biological effects of S1P depend on receptor subtype, cellular context, and interactions with inflammatory pathways [32, 37, 38]. These mechanisms further support the potential value of longitudinal S1P monitoring beyond a single baseline measurement.

Dynamic changes in several biomarkers, including lactate, CRP, PCT, and cytokines, have also been investigated in sepsis [7, 39]. In our cohort, IL-6 concentrations were also independently associated with mortality. However, individual biomarkers may reflect distinct aspects of sepsis pathophysiology. Therefore, combining biomarkers, including longitudinal S1P, may provide complementary prognostic information [34]. Importantly, research on S1P in sepsis remains in its infancy, with existing cohorts limited by small sample sizes and substantial heterogeneity. Future studies should validate these findings in larger, multicenter cohorts with standardized serial S1P measurements.

Several limitations should be acknowledged. First, this was a single-center study with a limited number of mortality events, which may have affected the precision and stability of the estimates. Second, recruitment was restricted to the ED, where patients may have less severe illness and lower mortality rates than those admitted to the ICU, limiting the generalizability of our findings to other clinical settings [40]. Third, residual confounding related to S1P carriers and treatments, including fluid resuscitation and albumin administration, cannot be excluded. Finally, the findings were not validated in an independent cohort, limiting their reproducibility and broader applicability.

## Conclusions

Baseline plasma S1P concentrations were lower in patients with sepsis. More importantly, declining S1P concentrations over time were associated with higher mortality, whereas baseline levels alone were not. These findings suggest that dynamic changes in S1P may provide clinically relevant information beyond a single baseline measurement.

## Data Availability

All data produced in the present study are available upon reasonable request to the authors

## List of abbreviations

ApoM: Apolipoprotein M
AUC: Area under the receiver operating characteristic curve
BMI: Body mass index
CCI: Charlson comorbidity index
CI: Confidence interval
CRP: C-reactive protein
ED: Emergency department
HDL: High-density lipoprotein
HR: Hazard ratio
ICU: Intensive care unit
IL-6: Interleukin-6
IQR: Interquartile range
PCT: Procalcitonin
S1P: Sphingosine-1-phosphate
S1PR: Sphingosine-1-phosphate receptor
SD: Standard deviation
SMD: Standardized mean difference
SOFA: Sequential Organ Failure Assessment

## Declarations

### Ethics approval and consent to participate

The clinical study was approved by the Taipei Medical University Institutional Review Board (TMU-JIRB No. N202202022), and was conducted in accordance with the Declaration of Helsinki. Written informed consent was obtained from all participants or from their legally authorized representatives.

### Availability of data and materials

The datasets used during the current study are available from the corresponding author upon reasonable request.

### Consent for publication

Not applicable.

### Authors’ contributions

B.V.T., C.L.C., and T.N.N. contributed to study conception, data curation, formal analysis, investigation, and drafting of the manuscript. Y.C.L. and K.C.H. contributed to investigation and data curation. C.W.H. contributed to methodology and manuscript review and editing. S.C.H. supervised the study, contributed to methodology, and reviewed and edited the manuscript. All authors reviewed and approved the final manuscript.

## Acknowledgements

The authors thank all participants who took part in this study and the clinical staff of the emergency department, Taipei Municipal Wanfang Hospital, for their support in patient recruitment and sample collection.

## References

1. Gray AP, Chung E, Hsu RL, Araki DT, Gershberg Hayoon A, Davis Weaver N, et al. (2025) Global, regional, and national sepsis incidence and mortality, 1990–2021: a systematic analysis. Lancet Glob Health 13(12):e2013-e26. 10.1016/S2214-109X(25)00356-0

2. Meyer NJ, Prescott HC (2024) Sepsis and septic shock. N Engl J Med 391(22):2133–46. doi:10.1056/NEJMra2403213

3. Singer M, Angus DC, Annane D, Bauer M, Kalil AC, Klompas M, et al. (2026) Sepsis. Lancet 407(10535):1276–88. 10.1016/S0140-6736(25)02422-5

4. Kumar A, Roberts D, Wood KE, Light B, Parrillo JE, Sharma S, et al. (2006) Duration of hypotension before initiation of effective antimicrobial therapy is the critical determinant of survival in human septic shock. Crit Care Med 34(6):1589–96. 10.1097/01.Ccm.0000217961.75225.E9

5. Seymour CW, Gesten F, Prescott HC, Friedrich ME, Iwashyna TJ, Phillips GS, et al. (2017) Time to treatment and mortality during mandated emergency care for sepsis. N Engl J Med 376(23):2235–44. doi:10.1056/NEJMoa1703058

6. Singer M, Deutschman CS, Seymour CW, Shankar-Hari M, Annane D, Bauer M, et al. (2016) The third international consensus definitions for sepsis and septic shock (sepsis-3). Jama 315(8):801–10. 10.1001/jama.2016.0287

7. Pierrakos C, Velissaris D, Bisdorff M, Marshall JC, Vincent JL (2020) Biomarkers of sepsis: time for a reappraisal. Crit Care 24(1):287. 10.1186/s13054-020-02993-5

8. Venkataraman K, Lee YM, Michaud J, Thangada S, Ai Y, Bonkovsky HL, et al. (2008) Vascular endothelium as a contributor of plasma sphingosine 1-phosphate. Circ Res 102(6):669–76. 10.1161/circresaha.107.165845

9. Venkataraman K, Lee YM, Michaud J, Thangada S, Ai Y, Bonkovsky HL, et al. (2008) Vascular endothelium as a contributor of plasma sphingosine 1-phosphate. Circ Res 102(6):669–76. 10.1161/CIRCRESAHA.107.165845

10. Ksiazek M, Chacinska M, Chabowski A, Baranowski M (2015) Sources, metabolism, and regulation of circulating sphingosine-1-phosphate. J Lipid Res 56(7):1271–81. 10.1194/jlr.R059543

11. Cartier A, Hla T (2019) Sphingosine 1-phosphate: Lipid signaling in pathology and therapy. Science 366(6463). 10.1126/science.aar5551

12. Christoffersen C, Nielsen LB (2013) Apolipoprotein M: bridging HDL and endothelial function. Curr Opin Lipidol 24(4):295–300. 10.1097/MOL.0b013e328361f6ad

13. Li Y, Zhou J, Qiu J, Huang Z, Wang W, Wu P, et al. (2020) Berberine reduces gut-vascular barrier permeability via modulation of ApoM/S1P pathway in a model of polymicrobial sepsis. Life Sci 261:118460. 10.1016/j.lfs.2020.118460

14. Proia RL, Hla T (2015) Emerging biology of sphingosine-1-phosphate: its role in pathogenesis and therapy. J Clin Invest 125(4):1379–87. 10.1172/jci76369

15. Piotti A, Novelli D, Meessen JMTA, Ferlicca D, Coppolecchia S, Marino A, et al. (2021) Endothelial damage in septic shock patients as evidenced by circulating syndecan-1, sphingosine-1-phosphate and soluble VE-cadherin: a substudy of ALBIOS. Crit Care 25(1). 10.1186/s13054-021-03545-1

16. Winkler MS, Nierhaus A, Holzmann M, Mudersbach E, Bauer A, Robbe L, et al. (2015) Decreased serum concentrations of sphingosine-1-phosphate in sepsis. Crit Care 19:372. 10.1186/s13054-015-1089-0

17. Winkler MS, Märtz KB, Nierhaus A, Daum G, Schwedhelm E, Kluge S, et al. (2019) Loss of sphingosine 1-phosphate (S1P) in septic shock is predominantly caused by decreased levels of high-density lipoproteins (HDL). J Intensive Care 7:23. 10.1186/s40560-019-0376-2

18. Wu XL, Hou JC, Li H, Xie GH, Zhang XL, Zheng JG, et al. (2019) Inverse correlation between plasma sphingosine-1-phosphate and ceramide concentrations in septic patients and their utility in predicting mortality. Shock 51(6):718–24. 10.1097/SHK.0000000000001229

19. Chang YJ, Yoo HJ, Kim SJ, Lee KH, Lim CM, Hong SB, et al. (2023) A targeted metabolomics approach for sepsis-induced ARDS and its subphenotypes. Crit Care 27(1):263. 10.1186/s13054-023-04552-0

20. Frej C, Linder A, Happonen KE, Taylor FB, Lupu F, Dahlbäck B (2016) Sphingosine 1-phosphate and its carrier apolipoprotein M in human sepsis and in Escherichia coli sepsis in baboons. J Cell Mol Med 20(6):1170–81. 10.1111/jcmm.12831

21. Kurano M, Tsuneyama K, Morimoto Y, Shimizu T, Jona M, Kassai H, et al. (2018) Apolipoprotein M protects lipopolysaccharide-treated mice from death and organ injury. Thromb Haemost 118(6):1021–35. 10.1055/s-0038-1641750

22. Pyne NJ, Pyne S (2010) Sphingosine 1-phosphate and cancer. Nat Rev Cancer 10(7):489–503. 10.1038/nrc2875

23. Fakhr Y, Brindley DN, Hemmings DG (2021) Physiological and pathological functions of sphingolipids in pregnancy. Cell Signal 85:110041. 10.1016/j.cellsig.2021.110041

24. Evans L, Rhodes A, Alhazzani W, Antonelli M, Coopersmith CM, French C, et al. (2021) Surviving sepsis campaign: international guidelines for management of sepsis and septic shock 2021. Intensive Care Med 47(11):1181–247. 10.1007/s00134-021-06506-y

25. Coldewey SM, Benetti E, Collino M, Pfeilschifter J, Sponholz C, Bauer M, et al. (2016) Elevation of serum sphingosine-1-phosphate attenuates impaired cardiac function in experimental sepsis. Sci Rep 6:27594. 10.1038/srep27594

26. Moritz E, Wegner D, Groß S, Bahls M, Dörr M, Felix SB, et al. (2017) Data on subgroup specific baseline characteristics and serum sphingosine-1-phosphate concentrations in the Study of Health in Pomerania. Data Brief 12:46–50. 10.1016/j.dib.2017.03.019

27. Daum G, Winkler M, Moritz E, Müller T, Geffken M, von Lucadou M, et al. (2020) Determinants of serum- and plasma sphingosine-1-phosphate concentrations in a healthy study group. TH Open 4(1):e12–e9. 10.1055/s-0040-1701205

28. Soltau I, Mudersbach E, Geissen M, Schwedhelm E, Winkler MS, Geffken M, et al. (2016) Serum-sphingosine-1-phosphate concentrations are inversely associated with atherosclerotic diseases in humans. PLoS One 11(12):e0168302. 10.1371/journal.pone.0168302

29. Johansson PI, Henriksen HH, Karvelsson ST, Rolfsson Ó, Schønemann-Lund M, Bestle MH, et al. (2024) LASSO regression shows histidine and sphingosine 1 phosphate are linked to both sepsis mortality and endothelial damage. Eur J Med Res 29(1):71. 10.1186/s40001-023-01612-7

30. Seidita I, Ziegler A, Qalaj A, Winkler MS, Nierhaus A, Kluge S, et al. (2024) Rapid determination of sphingosine 1-phosphate association with carrier molecules by flow-induced dispersion analysis to predict sepsis outcome. iScience 27(11):111168. 10.1016/j.isci.2024.111168

31. Zhang Z, Reinikainen J, Adeleke KA, Pieterse ME, Groothuis-Oudshoorn CGM (2018) Time-varying covariates and coefficients in Cox regression models. Ann Transl Med 6(7):121. 10.21037/atm.2018.02.12

32. Ziegler AC, Müller T, Gräler MH (2021) Sphingosine 1-phosphate in sepsis and beyond: Its role in disease tolerance and host defense and the impact of carrier molecules. Cell Signal 78:109849. 10.1016/j.cellsig.2020.109849

33. Zeng Y, Adamson RH, Curry FR, Tarbell JM (2014) Sphingosine-1-phosphate protects endothelial glycocalyx by inhibiting syndecan-1 shedding. Am J Physiol Heart Circ Physiol 306(3):H363–72. 10.1152/ajpheart.00687.2013

34. Ziegler AC, Gräler MH (2021) Barrier maintenance by S1P during inflammation and sepsis. Tissue Barriers 9(4):1940069. 10.1080/21688370.2021.1940069

35. Juffermans NP, Radermacher P, Joffre J (2026) Vascular permeability and loss of glycocalix in sepsis: the role of fluid resuscitation. Intensive Care Med 52(3):558–61. 10.1007/s00134-026-08293-w

36. Winkler MS, Nierhaus A, Poppe A, Greiwe G, Gräler MH, Daum G (2017) Sphingosine-1-phosphate: a potential biomarker and therapeutic target for endothelial dysfunction and sepsis? Shock 47(6):666–72. 10.1097/shk.0000000000000814

37. Salminen AT, McCloskey MC, Ahmad SD, Romanick SS, Chen KH, Houlihan W, et al. (2022) Molecular mechanisms underlying the heterogeneous barrier responses of two primary endothelial cell types to sphingosine-1-phosphate. Eur J Cell Biol 101(3). 10.1016/j.ejcb.2022.151233

38. Mohammed S, Bindu A, Viswanathan A, Harikumar KB (2023) Sphingosine 1-phosphate signaling during infection and immunity. Prog Lipid Res 92:101251. 10.1016/j.plipres.2023.101251

39. Yende S, Kellum JA, Talisa VB, Peck Palmer OM, Chang CH, Filbin MR, et al. (2019) Long-term host immune response trajectories among hospitalized patients with sepsis. JAMA Netw Open 2(8):e198686. 10.1001/jamanetworkopen.2019.8686

40. Finkelsztein EJ, Jones DS, Ma KC, Pabón MA, Delgado T, Nakahira K, et al. (2017) Comparison of qSOFA and SIRS for predicting adverse outcomes of patients with suspicion of sepsis outside the intensive care unit. Crit Care 21(1):73. 10.1186/s13054-017-1658-5

