## Supplementary Material for "Dynamic changes in plasma sphingosine-1-phosphate concentrations and mortality in sepsis: a prospective cohort study in the emergency department"

**Table S1. Baseline clinical signs and biological parameters of patients with non-septic infection and sepsis**

| | Non-septic infection (n = 36) | Sepsis (n = 156) | p* |
|---|---|---|---|
| **Clinical signs** | | | |
| Heart rate, bpm | 91.1 ± 20.9 | 98.9 ± 24.9 | 0.108 |
| Mean blood pressure, mmHg | 103.5 ± 23.5 | 95.2 ± 20.8 | 0.051 |
| Respiratory rate, bpm | 18.9 ± 4.7 | 21.1 ± 5.5 | **0.043** |
| Temperature, ºC | 36.8 ± 0.7 | 37.5 ± 1.2 | **0.003** |
| $SpO_2$, % | 96.5 ± 5.1 | 94.4 ± 6.4 | 0.097 |
| Urine output, mL/kg/min | 0.65 ± 0.40 | 0.54 ± 0.39 | 0.158 |
| **Baseline laboratory data** | | | |
| White blood cells, $\times 10^9$/L | 10.6 ± 4.5 | 11.9 ± 5.5 | 0.193 |
| Neutrophil, % | 69.2 (62.3–78.6) | 84.0 (76.1–90.0) | **<0.001** |
| Creatinine, mg/dL | 1.00 (0.77–1.25) | 1.08 (0.85–1.62) | 0.163 |
| C-reactive protein, mg/L | 2.5 (0.5–7.2) | 11.4 (5.0–18.3) | **<0.001** |
| Lactate, mmol/L | 1.1 (0.9–1.7) | 2.0 (1.4–3.1) | 0.059 |
| Platelet, $\times 10^9$/L | 248 (186–326) | 206 (163–265) | 0.062 |
| Albumin (g/L) | 3.54 ± 0.32 | 3.09 ± 0.54 | **0.032** |
| IL-6, pg/mL | 22.5 (9.0–64.1) | 101.5 (29.8–351.6) | **<0.001** |
| S1P, ng/mL | 129.0 (68.0–307.2) | 82.0 (48.0–141.0) | **<0.001** |
| SOFA score | 1.27 ± 1.55 | 3.40 ± 2.03 | **<0.001** |

**Footnotes:** $SpO_2$, peripheral oxygen saturation; IL-6, interleukin-6; S1P, sphingosine-1-phosphate; SOFA, Sequential Organ Failure Assessment; * p-values for comparisons between two groups (Chi-square test for categorical variables and independent-samples t-test or Mann–Whitney U test for continuous variables).

**Table S2. Comparison of baseline characteristics between sepsis and septic shock patients**

| | Sepsis (n = 130) | Septic shock (n = 26) | p |
|---|---|---|---|
| Age, years | 69.2 ± 16.8 | 77.1 ± 14.1 | **0.026** |
| Male, n (%) | 75 (57.7) | 15 (57.7) | 1.000 |
| Body weight, kg | 62.7 ± 17.7 | 60.1 ± 16.9 | 0.477 |
| **Comorbidity, n (%)** | | | |
| Heart failure | 16 (12.3) | 5 (19.2) | 0.529 |
| CKD | 26 (20.0) | 13 (50.0) | **0.003** |
| Diabetes | 42 (32.3) | 6 (23.1) | 0.485 |
| Hypertension | 77 (59.2) | 12 (46.2) | 0.311 |
| Hyperlipidemia | 10 (7.7) | 1 (3.8) | 0.780 |
| CCI, points | 2.49 ± 1.78 | 3.19 ± 2.02 | 0.107 |
| **Clinical signs** | | | |
| Heart rate, bpm | 99.2 ± 23.7 | 97.6 ± 30.6 | 0.765 |
| MAP, mmHg | 97.3 ± 20.7 | 84.4 ± 17.7 | **0.003** |
| Respiratory rate, bpm | 20.6 ± 5.3 | 23.5 ± 6.2 | **0.014** |
| Temperature, ºC | 37.5 ± 1.0 | 37.5 ± 1.7 | 0.726 |
| $SpO_2$, % | 95.1 ± 4.7 | 91.3 ± 11.3 | **0.006** |
| Urine output, mL/kg/min | 0.59 ± 0.38 | 0.3 ± 0.3 | **<0.001** |
| SOFA score | 2.99 ± 1.57 | 5.46 ± 2.77 | **<0.001** |
| **Laboratory data** | | | |
| Creatinine (mg/dL) | 1.0 (0.81–1.5) | 1.72 (1.1–2.4) | **0.002** |
| CRP (mg/L) | 11.1 (5.0–17.8) | 13.7 (5.8–19.5) | 0.266 |
| Lactate (mmol/L) | 1.8 (1.3–2.9) | 2.7 (1.6–3.3) | 0.056 |
| Albumin (g/L) | 3.2 ± 0.6 | 2.8 ± 0.4 | **0.005** |
| IL-6 (pg/mL) | 83.6 (21.1–261.3) | 237.0 (126.5–653.9) | **<0.001** |
| S1P (ng/mL) | 82.5 (51.8–144.5) | 79.0 (38.4–128.4) | 0.447 |
| Positive blood culture, n (%) | 38 (29.2) | 14 (53.8) | **0.028** |
| ICU admission rate, n (%) | 4 (3.1) | 14 (53.8) | **<0.001** |
| **Outcomes** | | | |
| Hospital length of stay, days | 10.0 (8.0–14.0) | 11.0 (9.0–17.3) | 0.326 |
| 60-day mortality, n (%) | 8 (6.2) | 14 (53.8) | **<0.001** |

**Abbreviations:** CKD, chronic kidney disease; CCI, Charlson Comorbidity Index; MAP, mean arterial pressure; $SpO_2$, peripheral oxygen saturation; SOFA, Sequential Organ Failure Assessment; CRP, C-reactive protein; IL-6, interleukin-6; S1P, sphingosine-1-phosphate; ICU, intensive care unit.

**Table S3. Correlations between baseline S1P concentrations and clinical parameters in the sepsis cohort**

| | n | Spearman rho | p* |
|---|---|---|---|
| White blood cells, K/μL | 156 | 0.041 | 0.611 |
| Neutrophil (%) | 156 | 0.135 | 0.094 |
| Hemoglobin, g/dL | 156 | −0.051 | 0.529 |
| Platelet, K/μL | 156 | −0.039 | 0.631 |
| CRP, mg/L | 156 | 0.082 | 0.312 |
| Lactate, mmol/L | 124 | −0.050 | 0.641 |
| Creatinine, mg/dL | 156 | 0.045 | 0.576 |
| Albumin, g/L | 105 | 0.084 | 0.514 |
| Glucose, mg/dL | 149 | 0.091 | 0.271 |
| IL-6, pg/mL | 154 | 0.250 | **0.002** |
| CCI | 156 | −0.128 | 0.098 |
| SOFA, points | 156 | −0.166 | **0.038** |

* Spearman rank test. CRP, C-reactive protein; IL-6, interleukin-6; CCI, Charlson Comorbidity Index; SOFA, Sequential Organ Failure Assessment.

**Table S4. Baseline covariates stratified by S1P trajectory groups after propensity score matching**

| | Ascending (n = 50) | Descending (n = 50) | SMD |
|---|---|---|---|
| Age, years | 72.7 ± 16.8 | 69.8 ± 16.6 | 0.129 |
| Male, n (%) | 30 (60.0) | 32 (64.0) | 0.082 |
| Body weight, kg | 62.0 ± 17.0 | 62.4 ± 20.3 | 0.022 |
| CCI, points | 2.5 ± 1.5 | 2.2 ± 1.4 | 0.118 |
| SOFA score, points | 3.2 ± 1.9 | 3.25 ± 1.7 | 0.011 |
| IL-6 (pg/mL) | 258.2 ± 299.1 | 251.9 ± 357.8 | 0.019 |

**Footnotes:** S1P, sphingosine-1-phosphate; SMD, standardized mean difference; CCI, Charlson Comorbidity Index; SOFA, Sequential Organ Failure Assessment; IL-6, interleukin-6.
